# Characterising severe traumatic brain injury from fresh cerebral biopsy in living patients

**DOI:** 10.1101/2021.10.27.21265411

**Authors:** Ping K. Yip, Shumaila Hasan, Christopher E.G. Uff

**Author notes:** Correspondence to: Drs Ping K. Yip and Chris E. Uff, Dr Ping K. Yip, Queen Mary University of London, Blizard Institute, Centre for Neuroscience, Surgery & Trauma, 4 Newark St, London E1 2AT, U.K., Dr Chris E. Uff, Department of Neurosurgery, Royal London Hospital, Whitechapel, London, E1 1FR, U.K.

## Abstract

Traumatic brain injury (TBI) is one of the most complex disorder in the most complex organ in the body and current classifications of mild, moderate and severe often fail to capture this complexity. Although the mainstay of prognosis involves several clinical classification systems, these systems are confined to macroscopic analysis. Therefore, we aim to use immunohistochemical examination of fresh brain biopsy samples to study the cellular and molecular changes caused by severe TBI.

Twenty-five adult patients suffering severe TBI were recruited into the Severe Head Injury Brain Analysis (SHIBA) study. Fresh brain biopsies obtained prior intracranial pressure (ICP) monitor insertion or during craniotomy underwent immunohistochemical analysis using the neuronal marker (NeuN), dendritic marker (MAP2), vascular markers (claudin-5 and vWF), and neuroinflammation markers (Iba1 and P2Y12) to investigate the injury severity at microscopic level.

Obtaining brain biopsy from the twenty-five patients in the study did not cause any additional burden to patient’s standard care and there were no adverse effects. The mean (± SD) Glasgow Outcome Scale-Extended (GOS-E) 3-months after injury was 4.0 (± 2.7), consisting of 64% unfavourable outcomes and 36% favourable outcomes. Immunostaining of brain tissue revealed various qualitative changes resulting in neuronal injury, dendritic injury, neurovascular injury, and neuroinflammation, which we classified into 4 subgroups for each injury type using the newly devised Yip, Hasan and Uff (YHU) grading system. Using this grading scale, patients with a total YHU grade of ≥ 11, 100% (n=11) had a GOS-E of ≤ 4, including death (54.5%), vegetative state (18.2%) and severe disability (27.3%) 3-months post injury. In contrast, those with a YHU grade of ≤ 8, 100% (n=8) had a GOS-E of 5 or higher, indicating a favourable outcome. Linear regression showed a significant negative correlation between the GOS-E and neuronal injury (R^2^ =0.240, p= 0.0129), dendritic injury (R^2^ =0.430, p= 0.0004), neurovascular injury (R^2^ =0.538, p<0.0001), and neuroinflammation (R^2^ =0.361, p= 0.0015).

Brain biopsy in severe TBI is a simple and safe procedure. Furthermore, immunofluorescence staining enables classification of this heterogeneous patient population into various categories of injury severity based on the cellular and molecular pathophysiology according to the YHU grading system. This new grading scale should facilitate a more precise diagnosis, similar to what is currently standard in oncology, allowing earlier and more accurate prognosis than current grading systems, thereby adding to the arsenal of resources available to guide clinical decision making in treating patients with severe TBI.

## Introduction

Traumatic brain injury (TBI) affects millions of patients annually worldwide and many suffer severe disability and death ^1,2^. When head injury occurs, the brain is subject to a multitude of forces, many unknown and generally chaotic, resulting in a plethora of injuries (epidural, subdural, subarachnoid and intracerebral bleeding, traumatic diffuse axonal injury, and both coup and contre-coup injury patterns) some of which are predictable and some which are not. Known and unknown factors including age, handedness, pre-existing medical conditions, anatomical variants, a history of TBI, and genetic predisposition all influence the course of the disease, resulting in a massive variation in functional outcome stemming from seemingly similar injury mechanisms and clinical presentations.

Intensive care can support the unconscious patient and deep anaesthesia reduces the metabolic requirements of the brain allowing it to tolerate reduced blood flow and higher intracranial pressure (ICP). Surgery can reduce ICP by evacuating haematomata, draining cerebrospinal fluid (CSF) or decompressive craniectomy, all with the intent of mitigating secondary injury. However, no therapeutic agent has been shown to alter the course of the primary brain injury despite more than 50 trials of 31 therapies at a cost of $1.1 billion since 1993 ^3^.

TBI is usually classified as mild, moderate and severe by various parameters including clinical severity (including imaging), pathoanatomic type, outcome and prognosis. Outcome measures in clinical trials are generally dichotomised. These systems often fail to capture sufficient detail in this complex and varied disease which may explain the massive difference in outcomes observed in seemingly similar injuries ^4^.

While oncology has for years benefited from accurate diagnosis based on histological examination allowing arduous and unpleasant treatments to be confidently recommended or withheld, TBI does not benefit from anything comparable. In the early stages, prognosis is often uncertain, particularly with respect to whether any meaningful recovery will occur. Even if the likely outcome is thought to be a very poor neurological recovery, care is frequently continued for months in the hope that recovery will be better than expected. Furthermore, treatments such as decompressive craniectomy and tracheostomy may be undertaken without robust evidence.

Glasgow coma scale (GCS) has been a mainstay of classification and prognostication for more than 40 years ^5^, however factors such as intoxication and sedation are confounders ^6^. Furthermore, it is possible that the universal failure of therapeutic agents, all of which showed promise in pre-clinical and/or phase I and II clinical trials, is in part due to inaccurate classification systems which lack adequate resolution resulting in dilution of results ^7^.

Medical imaging, while currently the cornerstone of TBI diagnosis, is as yet unable to detect important microscopic cellular changes due to a lack of spatial resolution. TBI can lead to various types of pathophysiological events such as neuronal injury, dendritic injury, neurovascular disruption, and neuroinflammation, ^8-10^. These cellular and molecular changes occur at microscopic levels and by the time clinical deterioration such as raised ICP, pupillary abnormalities or neuroimaging changes occur, it is often too late for effective treatment.

A better method of identifying the pathophysiology of TBI at the microscopic level is therefore required. Immunohistochemistry is a popular technique used universally in most scientific fields. During the past decade, the knowledge of neuronal injury, dendritic injury, neurovascular injury and neuroinflammation has vastly increased based *in vitro, in vivo* and post-mortem studies. For example, neuronal injury has been studied using neuronal nuclear protein (NeuN) in TBI ^8,10,11^, and dendritic injury using microtubule-associated protein-2 (MAP2) in rodent cortical injury ^12^. Neurovascular injury in TBI has been studied using claudin-5 and von Willebrand factor (vWF) ^13^. Neuroinflammation in traumatic CNS injuries, particularly in microglia has been studied using ionized calcium-binding adaptor molecule 1 (Iba1) and/or P2Y12 ^10,14^.

Brain biopsy remains the gold standard of diagnosis in neuro-oncology and has been used in other conditions including presenile patients ^15,16^ and normal pressure hydrocephalus ^17^. A single study using the technique of brain biopsy prior ICP monitor insertion in severe TBI demonstrated the safety of the procedure ^18^. Thus, the aims of this study were to affirm the safety of the procedure in severe TBI, and to determine the various pathophysiological responses after TBI at the microscopic level using immunohistochemistry (IHC) in fresh brain biopsies. The study was able to classify certain groups of brain injury pathologies that may be able to diagnose and prognosticate severe TBI patients based on the expression of a variety of immunostaining markers.

## Materials and methods

Ethical approval was granted by the London-Camden & Kings Cross Research Ethics Committee (REC ref: 20/LO/0074) and this application was supported by a patient and public involvement project where >90% of participants were supportive of brain biopsy in TBI ^19^. All patients were putatively unconscious at the time of enrolment and in line with the declaration of Helsinki, the 2005 UK Mental Capacity Act and the REC approved protocol, the next-of-kin were asked to act as a personal consultee, or if none were available, a senior doctor was asked to act as an Independent Healthcare Professional (IHP) consultee. Twenty-five consecutively eligible severe TBI patients were recruited at the Royal London Hospital between July 2020 to May 2021. Next of kin gave primary assent in 2/25 patients. In the remainder an IHP consultee was used. Two patients recovered to give their own consent prior to discharge and in the remainder of cases the next of kin subsequently gave consent, except in one case where no next of kin was identified and the patient had not regained capacity 6 months after injury.

### Patients

Inclusion criteria comprised age ≥ 18 years and TBI requiring diagnostic or therapeutic breeching of the dura as part of standard care. Exclusion criteria included coagulopathy (point of care or laboratory international normalized ratio (INR) testing was performed in all patients), current use of anticoagulants or antiplatelet agents. Safety was assessed by follow up imaging performed as part of standard care (i.e. extra scans were not performed). All 25 patients were followed up for at least 3 months.

### Tissue collection and processing

Brain tissue was sampled opportunistically if the dura was opened for diagnostic (ICP monitoring) or therapeutic (craniotomy) purposes. When sampling during ICP monitor insertion, a Kocher’s point twist drill craniostomy was fashioned, generally on the right side, and the dura perforated to allow insertion of the intraparenchymal ICP transducer. Prior to insertion of the transducer, a biopsy needle was introduced to a depth of 1cm and a single core biopsy of brain tissue was taken. The ICP monitor was then inserted. This method was also employed for tissue sampling during insertion of an external ventricular drain. During craniotomy a single core biopsy was taken at a depth of 1cm from the superior frontal gyrus using a biopsy needle. The brain biopsy tissue was immediately placed into 10% formalin (Figure 1A). After at least 2 h, the sample was transferred into 20% sucrose in 0.1M phosphate-buffered solution and stored at 4°C until further processed.

**Figure 1.**
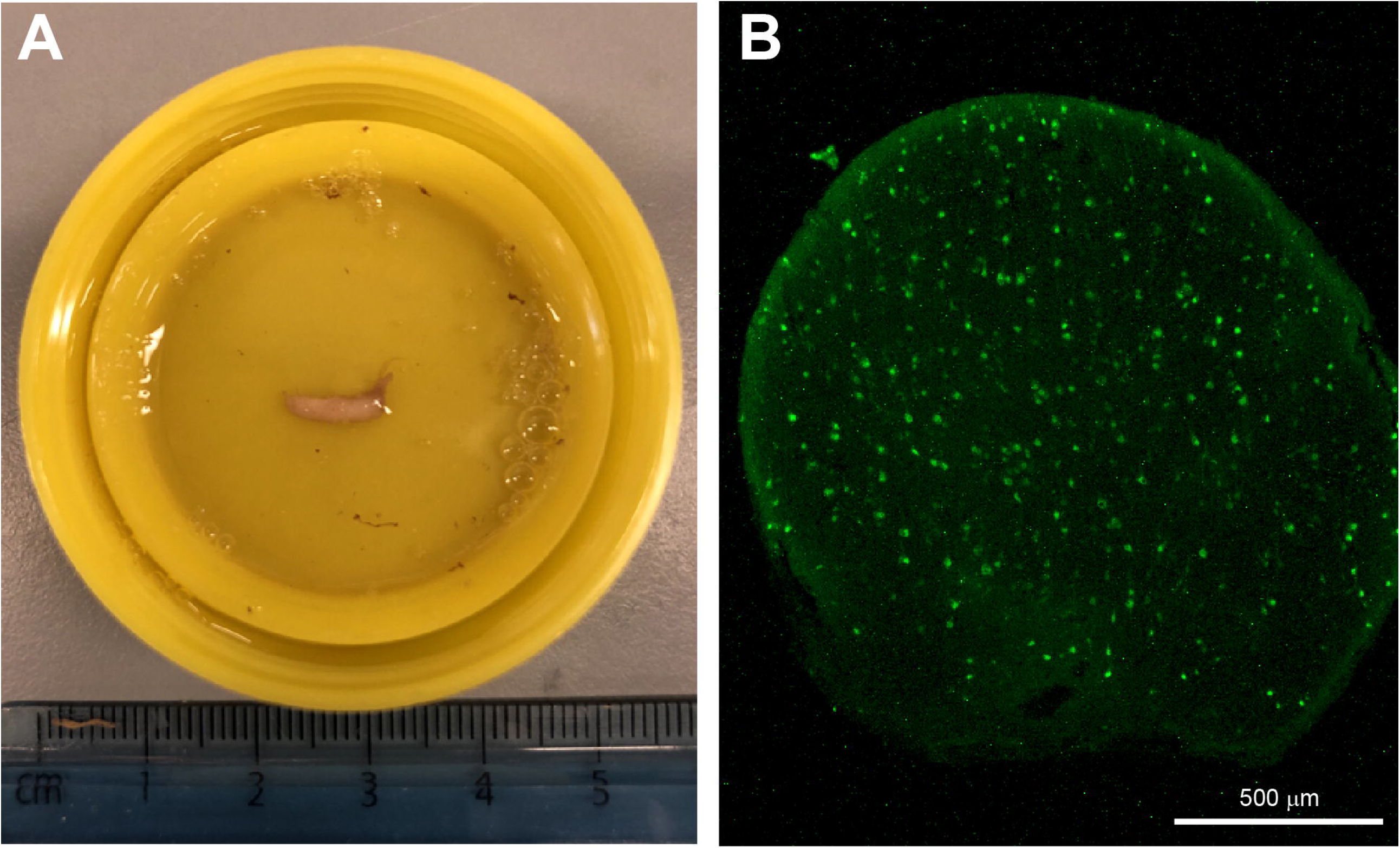
Cerebral biopsy sample. (**A**) An image of the cerebral biopsy was obtained from a severe TBI patient and fixed in 10% formalin. (**B**) A cross-section of the biopsy stained with the neuronal marker NeuN. Scale bar 500 μm.

If there was acute subdural haematoma present and if brain tissue or contusions were resected for therapeutic purposes, these samples were collected as additional specimen in 10% formalin and processed as above. Also, blood, cerebrospinal fluid (CSF) (if being drained therapeutically via an external ventricular drain (EVD)), saliva, urine and faeces were collected for 7 days when available. Analysis of these samples is beyond the remit of this study and will be reported separately.

### Immunohistochemistry

A small sample of the brain biopsy representing the dorsal region of the cerebral cortex was embedded into optimal cutting temperature (OCT) compound and then quickly frozen using dry ice. Using a freezing cryostat, the tissue was consecutively cut cross sectionally into 12μm thick sections and thaw-mounted onto microscope slides (Trajan Series 3). After at least one hour, the slides were further processed for immunohistochemistry as previously described (Yip et al., 2018). Briefly, the slides were washed with 3 × 5 minutes with 10mM phosphate-buffered saline (PBS), then incubated in antigen unmasking solution according to manufacturer’s protocol at 80°C for 30 min. After at least 10 minutes cooling at room temperature, the slides were washed with PBS for 3 × 5 minutes before being incubated with 2% skimmed milk (Marvel) for at least 30 minutes. Thereafter, the 2% skimmed milk was decarded and primary antibodies were applied. The primary antibodies used were: mouse anti-NeuN (1:500, #MAB377, Chemicon), mouse anti-MAP2 (1:500, #MAB3418, Merck Millipore, U.K.), mouse anti-claudin-5 (1:100, #35-2500, ThermoFisher), rabbit anti-vWF (1:200, #AB7356, Merck Millipore), rabbit anti-Iba1 (1:500, # 019-19741, Wako), rabbit anti-P2Y12 (1:400, #ANA55043A, AnaSpec, Cambridge Bioscience). Primary antibodies were allowed to incubate on the tissue sections overnight at room temperature in a sealed staining box. The following day, the slides were washed in PBS 3 × 5 minutes, then secondary antibodies were applied onto the sections and allow to incubate for two hours at room temperature. The secondary antibodies used were: donkey anti-rabbit Alexa Fluor® 488 or 594 (1:500, #A21206, #A21207, ThermoFisher), or donkey anti-mouse Alexa Fluor® 488 (1:500, #Ab150105, Abcam). Thereafter, the secondary antibodies were discarded, Hoechst (1:1000) was added for five minutes to counterstain the nuclei, then washed with PBS for 3 × 5 minutes before cover slipped using Vectashield® mounting medium (H-1000-10, Vector Labs). Negative controls were treated the same as the experimental slides but with the absence of the primary antibody (Figure 1C). The slides were visualised with a fluorescent microscope and images were captured at x20 and x40 magnification using a Kern ODC825 microscope digital camera and ToupView software.

### Data analysis and statistics

The stained brain sections were evaluated blind to the patient’s early clinical condition by PKY who has more than twenty years of histological experience with immunohistochemistry in traumatic CNS injuries, involving neuronal injury ^14,20^, dendritic injury ^21^, neurovascular injury ^9^, and neuroinflammation ^10,14^. The Glasgow Outcome Scale-Extended (GOS-E) at 3 months post injury was used to define the outcome of the patients as it is validated as a good indicator of severity, disability, cognitive dysfunction, and health in TBI patients ^22-24^. Linear regression analysis and graphical representation were carried out using GraphPad Prism v8. Data are expressed as mean ± standard deviation (SD). Statistical significance was determined when the p value <0.05.

## Results

### Demographics of patients

Of the 25 TBI patients recruited in the SHIBA study, 22 patients were male and 3 were female (Table 1). The mean (± SD) age of patients was 41.2 years (± 16.1) and the mean Glasgow Coma Scale (GCS) was 6.9 (± 3.6) at presentation (Table 1). Injury mechanisms included fall (44%), road traffic accident (32%), assault (12%), gunshot wound (8%), and train incident (4%).

**Table 1.** Summary clinical details of patients in this study.

| SHIBA code | TBI type | Age group (years) | Gender | Computerized tomography imaging | Biopsy time post injury (h) | trauma site | Biopsy site | Biopsy type | E | V | M | GCS | GOS-E (3 months) | Injury to death duration (day) |
| --- | --- | --- | --- | --- | --- | --- | --- | --- | --- | --- | --- | --- | --- | --- |
| 1 | RTC (motorcycle) | 21-25 | Male | Shallow ASDH / ICH / Contusion / tSAH / IVH | 146 | LF | LF | Craniotomy | 1 | 1 | 3 | 5 | 4 |  |
| 2 | RTC (motorcycle) | 21-25 | Male | Small EDH / Contusion / DAI | 5 | RF | RF | ICP | 1 | 1 | 5 | 7 | 7 |  |
| 3 | GSW | 16-20 | Male | Small ASDH / Contusion / tSAH | 1 | LT | RF | ICP | 2 | 2 | 5 | 9 | 5 |  |
| 4 | Assault | 40-45 | Male | Small contusions | 7 | LF | RF | ICP | 3 | 4 | 5 | 12 | 8 |  |
| 5 | Fall | 16-20 | Male | Left ASDH | 1 | L | RF | ICP | 1 | 1 | 1 | 3 | 7 |  |
| 6 | RTC (cyclist) | 36-40 | Male | Left ASDH / Diffuse contusions / Tight brain | 60 | LP | RF | ICP | 1 | 1 | 5 | 7 | 1 | 12 |
| 7 | Fall | 61-65 | Male | Left ASDH / LT contusions / All contrecoup | 8 | RP | LF | Craniotomy | 1 | 1 | 5 | 7 | 3 |  |
| 8 | Fall | 46-50 | Female | EDH and contusions | 1 | LP | RF | ICP | 1 | 1 | 1 | 3 | 8 |  |
| 9 | Fall | 16-20 | Male | DAI | 1 | RF | RF | ICP | 1 | 1 | 1 | 3 | 1 | 8 |
| 10 | Fall | 51-55 | Male | tSAH and contusions | 6 | OCC | RF | EVD | 3 | 2 | 5 | 10 | 7 |  |
| 11 | Assault | 31-35 | Male | tSAH | 3 | RF | RF | ICP | 1 | 1 | 4 | 6 | 8 |  |
| 12 | RTC (Pedestrian) | 56-60 | Male | ASDH and DAI | 3 | RP | LF | Craniotomy | 1 | 2 | 3 | 6 | 2 |  |
| 13 | Fall | 46-50 | Female | ASDH | 48 | RF | RF | Craniotomy | 4 | 2 | 5 | 11 | 7 |  |
| 14 | GSW | 21-25 | Male | GSW | 20 | LF | LF | Craniotomy | 4 | 4 | 6 | 14 | 3 |  |
| 15 | Fall | 76-80 | Male | IVH and contusions | 1 | OCC | RF | EVD | 1 | 2 | 5 | 8 | 1 | 5 |
| 16 | Fall | 31-35 | Male | tSAH and contusions | 4 | LP | RF | ICP | 3 | 4 | 6 | 13 | 3 |  |
| 17 | Fall | 56-60 | Male | ASDH | 4 | RP | LF | Craniotomy | 4 | 4 | 6 | 14 | 4 |  |
| 18 | Fall | 26-30 | Male | ASDH | 2 | BLF | RF | Craniotomy | 1 | 1 | 1 | 3 | 3 |  |
| 19 | RTC (Pedestrian) | 36-40 | Male | ASDH, contusions | 1 | LP | RF | Craniotomy | 1 | 1 | 2 | 4 | 1 | 5 |
| 20 | RTC (Pedestrian) | 31-35 | Male | Contusions and DAI | 3 | RF | RF | ICP | 1 | 1 | 2 | 4 | 2 |  |
| 21 | Assault | 51-55 | Female | Open depressed skull fracture | 1 | LF | LF | Craniotomy | 1 | 1 | 4 | 6 | 1 | 33 |
| 22 | Train accident | 56-60 | Male | DAI | 18 | LP | RF | ICP | 1 | 1 | 1 | 3 | 2 |  |
| 23 | RTC (Pedestrian) | 51-55 | Male | Contusions and DAI | 1 | OCC | RF | ICP | 1 | 2 | 3 | 6 | 1 | 2 |
| 24 | Fall | 56-60 | Male | ASDH / CSDH | 5 | RF | RF | Craniotomy | 1 | 1 | 3 | 5 | 4 |  |
| 25 | RTC (Pedestrian) | 46-50 | Male | ASDH, grade 3 DAI | 2 | RP | LF | Craniotomy | 1 | 1 | 1 | 3 | 1 | 4 |

Abbreviations: ASDH, acute subdural hematoma; BLF, bilateral frontal; CSDH, chronic subdural haematoma; DAI, diffused axonal injury; E, eye; EDH, epidural haematoma; EVD, external ventricular drainage; GCS, Glasgow coma scale; GOS-E, Glasgow outcome scale-extended; GSW, gunshot wound; ICH, intracerebral haemorrhage; ICP, intracranial pressure; IVH, intraventricular haemorrhage; L, left; LF, left frontal; LP, left parietal; LT, left temporal; M, motor; OCC, occipital; RF, right frontal; RP, right parietal; RTC, Road traffic accident; SHIBA, severe head injury brain analysis; TBI, traumatic brain injury; tSAH, traumatic subarachnoid haematoma; V, verbal.

At 3 months post injury, the mean (± SD) Glasgow Outcome Scale-Extended (GOS-E) was 4.0 (± 2.7), dichotomised between GOS-E score of 4 and 5, including 64 % vs 36% unfavourable vs favourable outcomes respectively (Table 1). Of the unfavourable outcome, GOS-E 1 (death) occurred in 28%, GOS-E 2 (vegetative state) in 12%, GOS-E 3 (lower severe disability) in 12% and GOS-E 4 (upper severe disability) in 12%. In contrast, of the favourable outcomes, GOS-E 5-6 (moderate disability) was seen in 4%, and GOS-E 7-8 (lower or upper good recovery) in 32%. Of the 7 patients who died, the mean (± SD) survival was 9.9 (± 10.7) days, ranging from 2 days to 33 days post injury (Table 1).

### Biopsy sampling

Fresh brain biopsies were obtained from the left (28%) or right (72%) superior frontal gyrus. Biopsy was performed prior to ICP monitor insertion in 48%, prior to EVD insertion in 8%, and at craniotomy in 44%. In 11 patients (44%) the biopsy was taken from a site remote to the primary (coup) or contrecoup injury. Mean (± SD) sampling time was at 14 h (± 31), with a mode of 1 h: 32% were carried out within 1 hour, 20% within 8 hours, 8% within 24 hours, and 12% greater than 48 hours after injury. We aimed to sample as soon as possible after injury and this range represent patients who suffered clinical deterioration while in hospital or who had a long lie prior to being found.

Twenty-four patients had a further CT scan within 24 hours of the biopsy as part of standard medical care. One patient recovered rapidly and there was no clinical indication for further imaging, although a delayed MRI scan showed no evidence of haemorrhage. Punctate haemorrhage was observed at the biopsy site in two patients (8%). In one patient, significant blossoming of frontal contusions extended into the biopsy site, but this was thought to be related to the primary injury rather than the biopsy.

### Immunohistochemistry

All biopsy samples collected were of sufficient size and quality to carry out the immunohistochemical analysis. Generally, the IHC procedures in this study were completed from sample collection to microscopic analysis within a 36-h time frame. However, it was possible to successfully complete the whole IHC procedure within 24-h (data not shown). To determine the severity of injury, immunofluorescence staining was used to classify the neuronal injury, dendritic injury, neurovascular injury, and neuroinflammation into 4 grades termed the Yip, Hasan and Uff (YHU) grading system. Grade I exhibited limited injury response in comparison to grade IV which had the most severe injury response. The YHU grading system was devised based on information from existing literature and personal knowledge (PKY).

### Neuronal injury

Neurones from non-injured brain immunostained with NeuN exhibited a large round or polygonal cell body with staining in both the nuclei and cytoplasm, and some proximal processes ^9,25,26^. However, neuronal atrophy, vacuoles in the cytoplasm, and neuronal cell loss were observed after injury ^27^.

According to the YHU grading system, grade I NeuN+ neurones with limited injury were observed in 8% of patients (Figure 2A, Table 2). Grade II containing a few neurones with small vacuoles in the cytoplasm was observed in 48% of patients (Figure 2B). Grade III consisting of several NeuN+ neurones with a dysmorphic appearance and/or large vacuolisation within the neuronal cytoplasm was observed in 24% of patients (Figure 2C). Grade IV consisting of a limited number of NeuN+ neurones with an atrophic appearance was observed in 20% of patients (Figure 2D). Overall, the mean (± SD) YHU grade for neuronal injury in this study cohort was 2.6 (± 0.9).

**Table 2.** The neuropathological examination of the fresh biopsy samples from the SHIBA patient cohort. The roman numerals in columns 2-5 represent the YHU grading scales presented in figures 2-5. Total YHU grade is a combination of all the grade scores of the 4 injury categories. Patient death or level of disability at 3 months post injury was defined by the GOS-E score.

| **SHIBA**<br>**Patient**<br>**No.** | **Neuronal**<br>**injury** | **Dendritic**<br>**injury** | **Neurovascular**<br>**injury** | **Neuro**<br>**inflammation** | **Total**<br>**YHU**<br>**grade** | **GOS-E** |
| --- | --- | --- | --- | --- | --- | --- |
| 1 | II | III | III | II | 10 | 4 |
| 2 | I | I | II | II | 6 | 7 |
| 3 | III | I | II | I | 7 | 5 |
| 4 | III | I | II | I | 7 | 8 |
| 5 | II | I | I | I | 5 | 7 |
| 6 | III | III | III | II | 11 | 1 |
| 7 | IV | IV | II | III | 13 | 3 |
| 8 | II | I | II | II | 7 | 8 |
| 9 | III | IV | IV | II | 13 | 1 |
| 10 | II | II | III | III | 10 | 7 |
| 11 | II | I | II | II | 7 | 8 |
| 12 | II | II | III | III | 10 | 2 |
| 13 | II | II | II | II | 8 | 7 |
| 14 | I | II | II | IV | 9 | 3 |
| 15 | II | I | III | III | 9 | 1 |
| 16 | II | II | II | II | 8 | 8 |
| 17 | II | III | IV | IV | 13 | 4 |
| 18 | II | II | III | II | 9 | 3 |
| 19 | IV | III | IV | III | 14 | 1 |
| 20 | IV | IV | IV | IV | 16 | 2 |
| 21 | III | II | III | IV | 12 | 1 |
| 22 | II | IV | IV | III | 13 | 2 |
| 23 | IV | IV | IV | IV | 16 | 1 |
| 24 | III | III | III | II | 11 | 4 |
| 25 | IV | IV | IV | IV | 16 | 1 |

**Figure 2.**
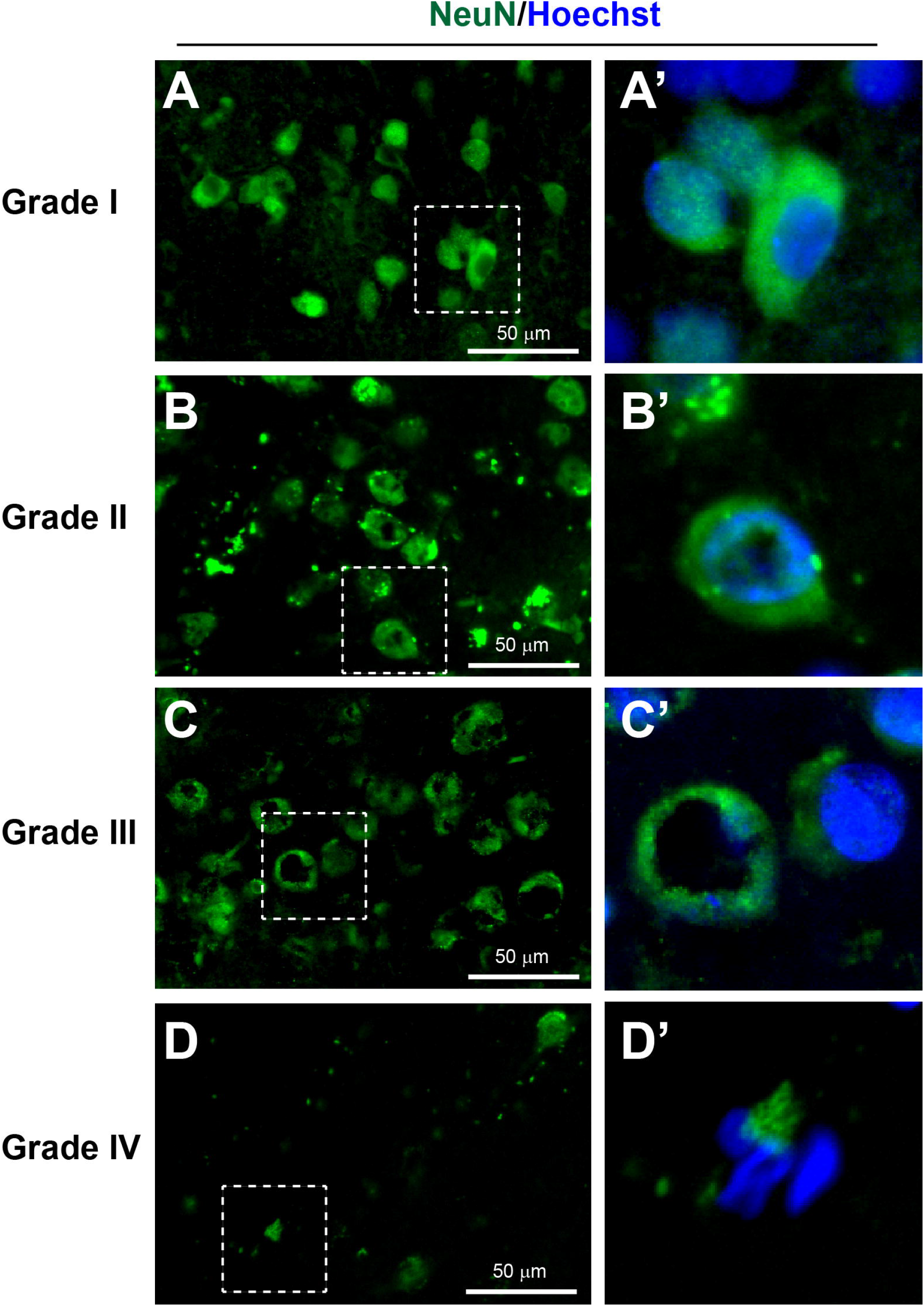
YHU grading system for the neuronal injury within the cerebral biopsy immunostained with NeuN (green) and Hoechst (nuclei, blue). (**A-A’**) Neurones with limited injury display large round/or polygonal cell bodies (grade I). (**B-B’**) Neurones with mild damage exhibit mild cellular shrinkage and/or small vacuolisation within the cytoplasm (grade II). (**C-C’**) Neurones with moderate damage will exhibit moderate cellular shrinkage and/or many with large cytoplasmic vacuolisations (grade III). (**D-D’**) Severe neuronal death indicated by atrophic cell bodies and few if any NeuN+ neurones (grade IV). (**A’-D’**) Enlarged images of the corresponding dashed boxes. Scale bar is 50 μm.

### Dendritic injury

Microtubule-associated protein 2 (MAP2) is a specific dendritic marker that has been shown to strongly immunostain dendrites and some nerve cell bodies, and weakly stain axons ^21,28^. Moreover, in tissue sections, healthy dendrites exhibit strong MAP immunostaining with smooth and continuous processes. However, dendrites after injury can develop swelling or a bead-like display termed dendritic beading ^29^.

According to the YHU grading system grade I consisting of MAP2+ dendrites with limited injury response was observed in 28% of patients (Figure 3A, Table 2). Grade II consisting of a few MAP2+ dendritic beadings was observed in 28% of patients (Figure 3B). Grade III consisting of extensive MAP2+ dendritic beading was observed in 20% of patients (Figure 3C). Grade IV consisting of limited MAP2+ dendritic staining with only the presence of a few strands of dendritic beading was observed in 24% of patients (Figure 3D). Overall, the average YHU grade for dendritic injury in this study cohort was 2.4 (± 1.2).

**Figure 3.**
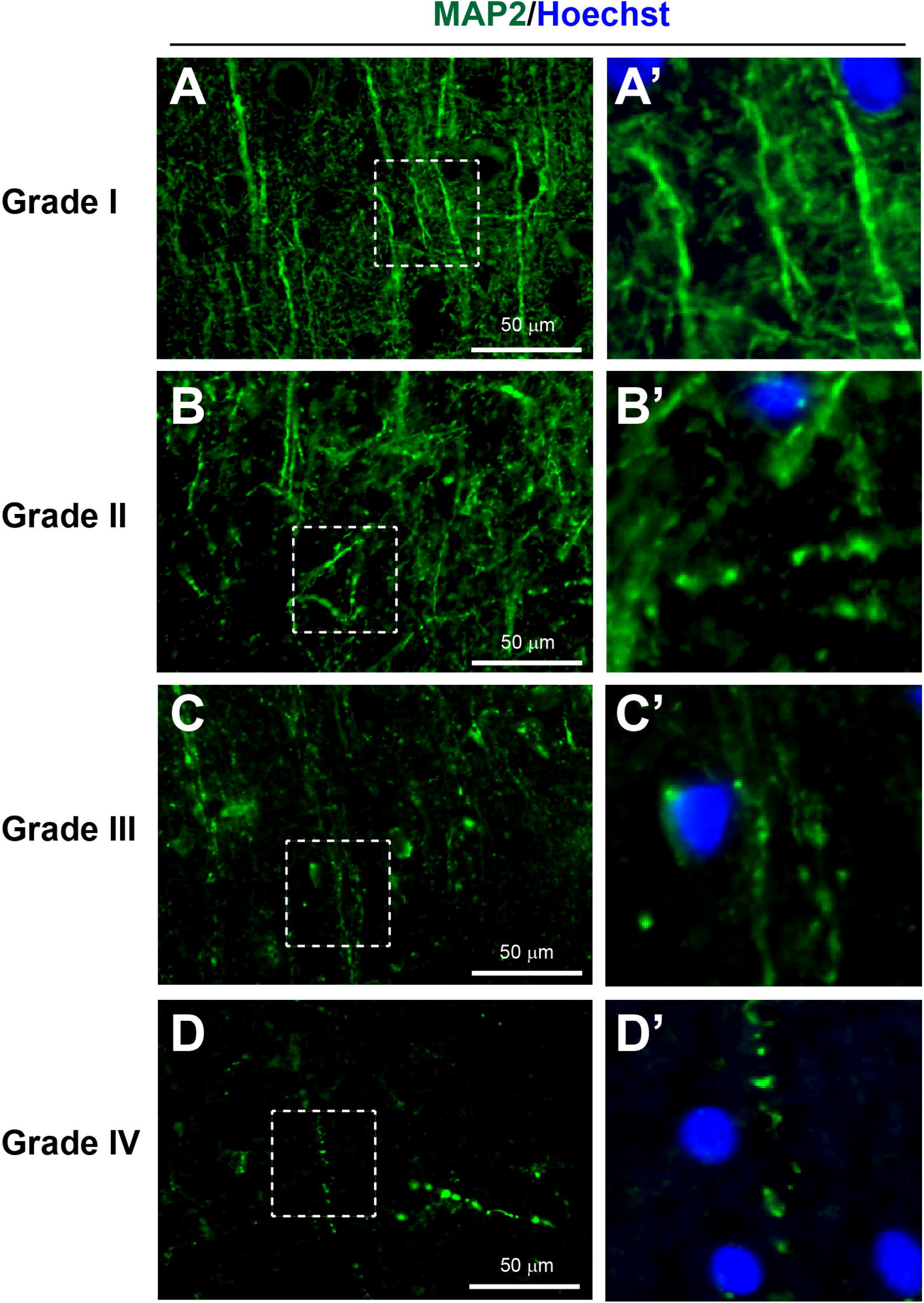
YHU grading system for the dendritic injury within the cerebral biopsy immunostained with MAP2 (green) and Hoechst (nuclei, blue). (**A-A’**) Dendrites with limited injury display strong continuous MAP2+ staining within some cell bodies and along the dendrites (grade I). (**B-B’**) Dendrites with mild damage exhibit reduced MAP2 staining and some signs of dendritic beading (grade II). (**C-C’**) Moderate dendritic injury exhibited moderate dendritic beading and some complete loss of dendrites (grade III). (**D-D’**) Severe dendritic injury is indicated by few if any dendrites present and only dendrites with beading present (grade IV). (**A’-D’**) Enlarged images of the corresponding dashed boxes. Scale bar is 50 μm.

### Neurovascular injury

The tight junction marker claudin-5 ^30^ and von Willebrand factor (vWF) that is present in endothelial cells ^31^ have shown healthy cerebral microvessels as long regular tubular shapes of approximately less than 10 μm in diameter. In microvascular damage after TBI, fragmented and/or absence of microvessels can be observed ^32^.

According to the YHU grading system, grade I consisting of several regular claudin-5+ and vWF+ long and intact tubular-shaped microvessels was observed in 4% of patients (Figure 4A, Table 2). Grade II consisting of many short microvessels was observed in 36% of patients (Figure 4B). Grade III consisting of extremely short segments of microvessels was observed in 32% of patients (Figure 4C). Grade IV consisting of irregular fragments or absence of any claudin-5+ and/or vWF+ microvessel staining was observed in 28% of patients (Figure 4D). Overall, the average YHU grade for neurovascular injury in this study cohort was 2.8 (± 0.9).

**Figure 4.**
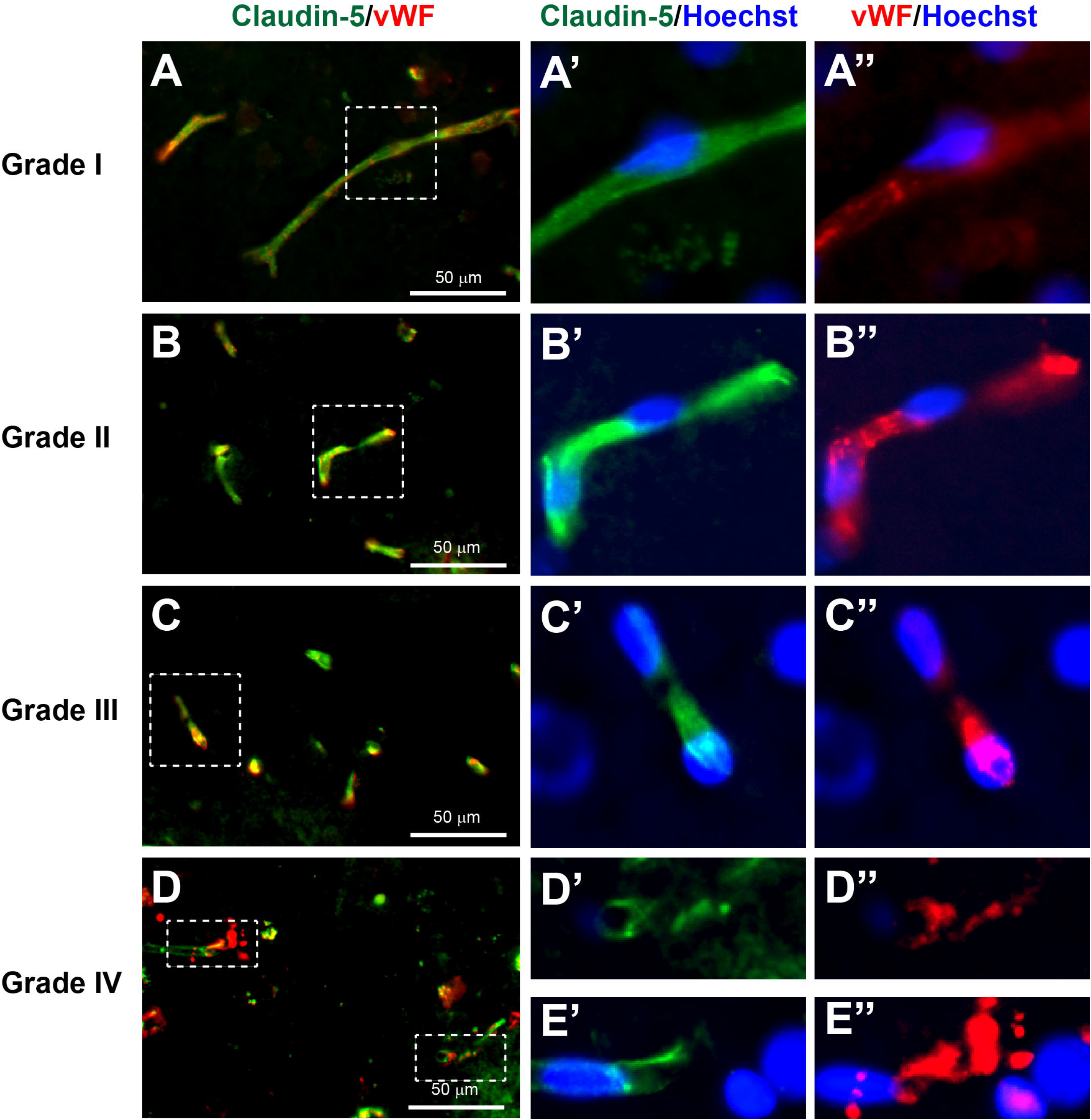
YHU grading system for the neurovascular injury within the cerebral biopsy immunostained with claudin-5 (green), von Willebrand factor (vWF, red), and Hoechst (nuclei, blue). (**A-A’’**) Microvessels with limited damage display moderate claudin-5 and vWF staining within long uninjured segments of microvessels (grade I). (**B-B’’**) Mild damaged microvessels exhibit stronger claudin-5 and vWF staining, with shorter microvessel segments (grade II). (**C-C’’**) Moderate damage to microvessels will exhibit predominantly very short segments (grade III). (**D-E’’**) Severe microvessel damage is indicated by few if any claudin-5 or vWF stained microvessels present and/or microvessels showed signs of ‘bursting’ morphology with large release of vWF bodies (grade IV). (**A’-E’’**) Enlarged images of the corresponding dashed boxes. Scale bar is 50 μm.

### Neuroinflammation

Immunohistochemical markers of microglia with Iba1 and P2Y12 have been widely used in previous studies of neuroinflammation ^10,14,33,34^. In healthy brain, microglia exhibit a ramified morphology with long thin processes and a small cell body. However, during neuroinflammation, the processes of microglia become thicker and shorter, and eventually exhibit an amoeboid-like shape with retraction of processes ^35^.

According to the YHU grading system. grade I consisting of predominantly Iba1+ and/or P2Y12+ microglia with a ramified morphology was observed in 12% of patients (Figure 5A, Table 2). Grade II consisting of many Iba1+ and/or P2Y12+ microglia with shorter and thicker processes was observed in 40% of patients (Figure 5B). Grade III consisting of predominantly Iba1+ and/or P2Y12+ amoeboid-like shape microglia with retracted processes was observed in 24% of patients (Figure 5C). Grade IV consisting of fragments and limited or absence of any Iba1+ and/or P2Y12+ microglia staining was observed in 24% of patients (Figure 5D). Overall, the average YHU grade for neuroinflammation in this study cohort was 2.6 (± 1.0).

**Figure 5.**
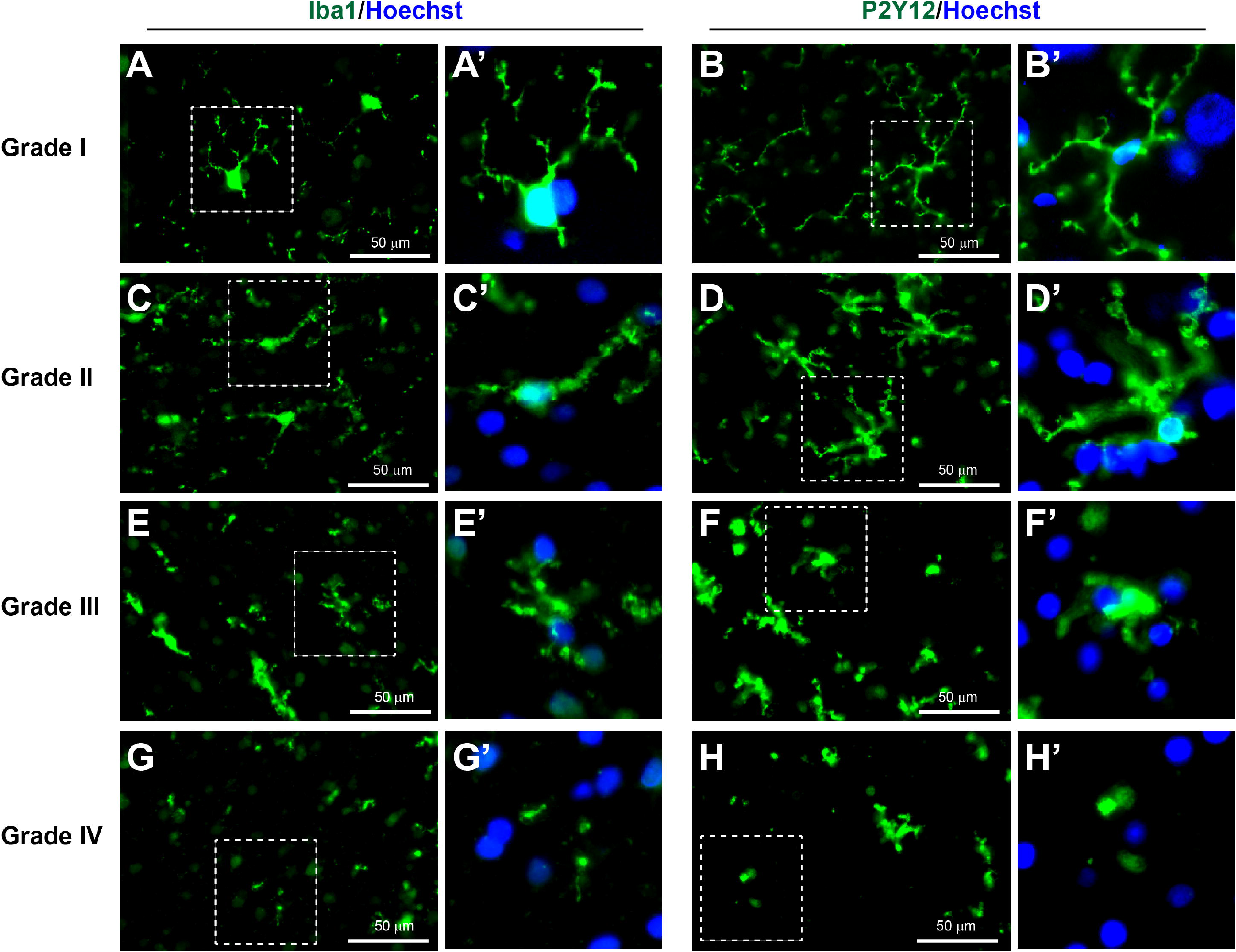
YHU grading system for the neuroinflammation within the cerebral biopsy immunostained with Iba1 (green, left panels), P2Y12 (green, right panels), and Hoechst (nuclei, blue). (**A-B’**) Microglia exposed to limited brain injury display a ramified appearance with long and thin processes and moderate expression of either iba1 and P2Y12 (grade I). (**C-D’**) Mild brain damage induced neuroinflammation with increase in microglial Iba1 and P2Y12 expression and thickening of processes (grade II). (**E-F’**) In moderate neuroinflammation, the microglia display towards an ameboid morphology with very short processes (grade III). (**G-H’**) In several brain injury, few if any intact microglia were present and signs of severe fragmentation occurred (grade IV). (**A’-H’**) Enlarged images of the corresponding dashed boxes. Scale bar is 50 μm.

### Diagnosis and Prognosis

The main objective for studying fresh brain tissue in TBI was to diagnose the severity of TBI based on the cellular and molecular changes that occur, and to correlate these with the clinical course and outcome with the intent of providing an early and accurate prognosis that may guide clinical decision making. Linear regression was carried out to compare GOS-E with the different YHU injury gradings. Linear regression between YHU grading of neuronal injury and GOS-E showed significant negative correlation (Figure 6A, R^2^ =0.240, p= 0.0129). Furthermore, linear regression between YHU grading of dendritic injury and GOS-E showed significant negative correlation (Figure 6B, R^2^ =0.430, p= 0.0004). Linear regression between YHU grading of neurovascular injury and GOS-E showed a significant negative correlation (Figure 6C, R^2^ =0.538, p<0.0001) which was less than the correlation between YHU grading of neuroinflammation and GOS-E (Figure 6D, R^2^ =0.361, p= 0.0015). Interestingly, linear regression between total YHU grading scores and presentation GCS was not significant (Figure 6E, R^2^ =0.073, p = 0.1913). However, linear regression between total YHU grading scores and GOS-E did show significant negative correlation (Figure 6F, R^2^ =0.584, p < 0.0001).

**Figure 6.**
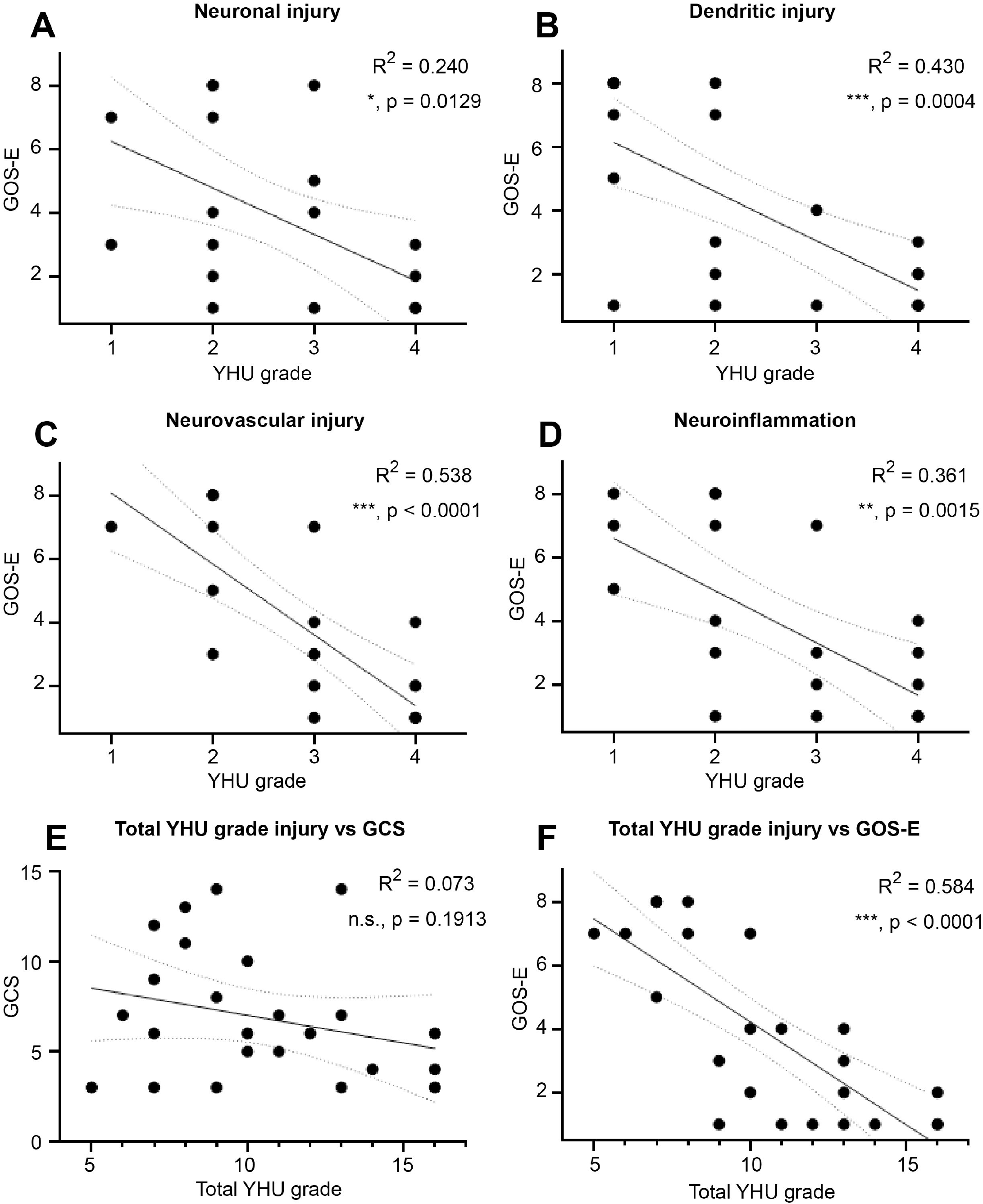
Linear regression analysis between the YHU grading system and GOS-E. The GOS-E was compared with the YHU grading for (**A**) neuronal injury, (**B**) dendritic injury, (**C**) neurovascular injury, (**D**) neuroinflammation. Combination of all YHU grading scores for neuronal injury, dendritic injury, neurovascular injury, and neuroinflammation of each patient and compare with (**E**) GCS and (**F**) GOS-E. n.s. = non-significant, * = p < 0.05, ** = p < 0.01, *** = p < 0.001.

Of the 10 patients with a GOS-E 1 (deceased) or 2 (vegetative state), 100% had a YHU grade III or IV in at least 2 injury types, and 80% had a YHU grade of III or IV in at least 3 injury types (Table 2). However, within the 7 patients with a GOS-E 3 or 4 (severe disability), 67% had a YHU grade III or IV in at least 2 injury types, and 50% had a YHU grade III or IV in at least 3 injury types (Table 2). In contrast, within the 8 patients with a GOS-E >5 (moderate disability to good recovery), 13% had a YHU grade III in at least 2 injury types, and 75% had only a YHU grade I or II in all injury types (Table 2). In summary, using the YHU grading system, patients with a total YHU grade of ≥ 11, 100% of the 11 patients had a GOS-E ≤ 4, which includes death (55%), vegetative state (18%) and severe disability (27%). In contrast, a total YHU grade of ≤ 8, 100% of the 7 patients had a GOS-E ≥ 5, indicating favourable outcome.

## Discussion

Currently, there are no effective treatments for severe TBI and to date, 31 large Phase III clinical trials have failed to exhibit any favourable findings ^3^. One possible reason may be the complex heterogeneity of TBI, especially within the severe TBI group is not detected by the simplistic mild, moderate and severe TBI classification systems based on clinical parameters which are used to validate these studies. In this study, we have demonstrated that using fresh brain biopsies taken at or soon after admission in 25 patients, severe TBI can be further subclassified using the YHU grading system into 4 grades of injury based on the patients’ severity of neuronal injury, dendritic injury, neurovascular injury and neuroinflammation using immunohistochemistry. These four parameters may yet yield further detail and clinical correlation in larger studies. In YHU grade I, the neurones, dendrites, microvessels and microglia exhibit a predominantly uninjured appearance. In YHU grade II, there are signs of mild injury to the neuronal cell bodies, dendrites, microvessels and activation of microglia. In YHU grade III, the neurones, dendrites, microvessels are significantly injured and microglia are extremely activated. In YHU grade IV, there is severe damage to neurones, dendrites, microvessels and microglia to the extent of absence. Patients with more YHU grade III and IV types of injury are more likely to have an unfavourable outcome compared to those with more YHU grade I and II types of injury who are more likely to have favourable outcome, and this information is potentially available as soon as 24 hours after injury.

Performing brain biopsy in living patients is not a new procedure. It has been conducted in more than 20 studies over 6 decades and fear of significant complications appears to be unwarranted ^16^. Although the safety of brain biopsy in severe TBI at the time of ICP bolt insertion has previously been demonstrated ^18^, establishing safety was an additional objective of this study, and as we have shown, in this population of 25 patients, a single core brain biopsy did not cause any significant or clinically detectable adverse effects. Single core biopsies were obtained to mitigate risk which is fewer than in some other studies, one which had a mean biopsy sample number of 8 in living presenile patients which also demonstrated no major adverse effects ^15^. Similar to this study, Castejon and colleagues collected cortical brain biopsies during craniotomy from 8 living TBI patients ranging from 1 to 35 days post injury and carried out traditional histological staining ^36^, successfully identifying the pathogenic mechanism of brain oedema. Brain biopsy is the cornerstone of neuro-oncological diagnosis, successfully providing a histological diagnosis with low morbidity and mortality ^37^, and with a greater than 90% accuracy of diagnosis of tumours and lesions in immunodeficient patients ^38^. We believe that the field of TBI research has the potential to follow a similar direction and progression to those recently made in brain tumours.

### Injury Markers

NeuN+ neuronal cells have been used in rodents to determine neurodegeneration after TBI ^10,25,39^. Therefore, it was not surprising to observe 92% of severe TBI patients in this study express some form of neuronal injury ranging from small vacuolization (YHU grade II) to major neuronal cell loss (YHU grade IV).

The loss of MAP2+ dendritic staining in YHU grade II - IV dendritic injury is in agreement with the loss of MAP2 staining in human post mortem brain samples from patients who died of hypoxia-ischemia ^28^ and within 3 hours after a lateral cortical impact injury in a rodent severe TBI model, immunohistochemistry demonstrated a substantial loss of MAP2 in the apical dendrites and dendritic arbors at the impact site ^12^. It has been shown *in vitro* that mechanical stretch injury to axons rather than dendrites causes dendritic beading, which can develop maximally within 5 minutes and last for up to 5 hours after injury ^29^. Interestingly, it has also been shown that dendritic beading can occur due to glucose toxicity ^40^ and oxygen-glucose deprivation ^41^, although it has been proposed that it is possible to have significant structural loss in dendritic arborisation without substantial neuronal loss. However, the critical role of spines on dendrites in synaptic transmission would be lost and significantly impaired neuronal function within the brain would result ^42^.

The enriched microvasculature in the CNS is tightly bound by the Blood Brain Barrier (BBB) to provide a physical and metabolic barrier for normal brain homeostasis ^43^. However, after TBI, there is increased vascular leakage due to disruption of the endothelial integrity ^44^. The damage and/or loss of claudin-5+ and/or vWF+ stained microvessels in the YHU grade II – IV neurovascular injury suggests that disruption and/or rupture of the neurovasculature had occurred in these patients. Therefore, the greater the damage to brain microvessels, the greater the breakdown of the BBB resulting in oedematous and/or hypoxic-ischemic brain injury ^36^.

Microglial activation has been the focus investigation into neuroinflammation, and potential therapies for TBI have attempted to reduce the microglial activation with the intent of limiting the injury and conferring neuroprotection ^10,45-47^. Therefore, this treatment strategy has the potential to benefit patients with YHU grade II – III neuroinflammation. However, in patients with YHU grade IV neuroinflammation, the detrimental effect after TBI is potentially not the activation of microglia, but rather the loss of microglial function that is important in normal brain function ^48^. This also applies to the loss of neurones, dendrites, microvasculature resulting in an inability to perform their normal roles, which may contribute to their unfavourable outcome.

Of the 10 patients who died or remained in persistent vegetative state, two patients exhibited a maximum of YHU grade III in two injury types, which was not in agreement with the other patients with a similar outcome. The reason for this discrepancy is not known, but potential contributory factors include age (the patient who died was within the age range of 76-80 years old) and injury pattern (the patient in vegetative state had a severe isolated injury to the pons).

The loss of any cell types such as neurones, glia cells and endothelial cells within microvessels is most likely due to tissue destruction caused by the primary brain injury. It was particularly surprising that this also occurred in brain biopsies that were taken from a location remote from the site of impact. In such severe injury, it seems unlikely that any potential therapy would benefit these patients, who accounted for 40% of this study. The inclusion of these patients may be an unknown potential confounding factor in the failure of clinical trials in severe TBI ^6^. Furthermore, the difference in some patients with neuronal injury, dendritic injury, neurovascular injury and neuroinflammation suggests that different types of treatment focussing on the predominant injury response is more likely to succeed. Therefore, early diagnosis and prognosis using this cellular and molecular grading system has the potential to provide a significant and accurate additional resource to not only guide patient care but to aid with future clinical trials.

### Strengths and limitations

This study has several strengths. The use of fresh brain biopsies obtained from living patients suffering TBI provides a precise and accurate analysis of selective cellular and molecular changes in the injured brain tissue. A very small sample of brain tissue collected as early as 1 h can successfully analyse neuronal injury, dendritic injury, neurovascular injury, and neuroinflammation caused by TBI. This patient cohort consisted of a large variety of head injury mechanisms representative of the variation of head injury types inflicted on the civilian population. The histologist (PKY) was initially blinded as to the early clinical course when providing the YHU grade for the immunohistological staining, so avoided any bias calls. The immediate transfer of the fresh brain samples into formalin enables the cellular and molecular changes to be minimised in comparison to post mortem samples which may take up to 2 days after death before tissue fixation ^28^ and that inadequate perfusion-fixation of a large tissue mass may be responsible for certain false positive pathological observations such as ‘dark’ neurones in the post mortem tissue ^36,49^.

This study also has several limitations. The 25 patients were all recruited at a single institution (The Royal London Hospital) and are a snapshot of the types of severe TBI that are observed. Patients taking anticoagulants, antiplatelet agents and known alcoholics with likely coagulopathy were excluded to reduce the potential risk of biopsy-induced intracerebral haemorrhage. Brain material from normal healthy control subjects was unavailable, so we were unable to provide a normal non-injured immunostaining expression for the markers used in this study. However, brain biopsy in healthy individuals is limited by ethical constraints. It is known that TBI is an extremely complex spectrum of disorders and the YHU grading scale is only an estimate of severity rather than a definitive scoring system. We believe however that this is the inception of a new era of immunohistochemical science in TBI and it is anticipated that the classification of severe TBI will continue to expand with more TBI-induced specific markers. While we do not presume to claim that it will eventually provide a definitive answer, we strongly believe that it will add significantly to the armamentarium of tools available to diagnose and prognosticate clinically, and to validate future clinical trials in TBI research.

Future potential applications to low- and middle-income countries are equally exciting as the biopsy and immunohistochemical staining may be performed at low cost. With standardized staining techniques and web-based upload of micrographs that can be interpreted by artificial intelligence algorithms, this and future grading systems will not just be in the remit of the first world.

## Conclusion

This was the first study to carry out immunohistochemical analysis on fresh brain biopsy from living severe TBI patients obtained as early as 1 h post TBI, and has shown that multipathological factors of varying severity of neuronal injury, dendritic injury, neurovascular injury and neuroinflammation can be classified into 4 subgroups using the YHU grading system. Furthermore, the YHU grading was shown to be significantly negatively correlated with the GOS-E at 3 months post injury. These findings have demonstrated that brain biopsy in TBI is safe and that cellular and molecular data could potentially be used to guide clinical decisions. Furthermore, this study suggests that there are certain TBI patients with extremely severe brain injury who are unlikely to benefit from any potential surgical and/or pharmacological treatment, and using the YHU classification, this information is available much earlier than by observing the patient’s clinical course. In summary, we propose that fresh brain biopsies should be used for precise and accurate diagnosis and prognosis of severe TBI, and should be recommended for use in future clinical trials to minimise the number of patients required to demonstrate the efficacy of a particular therapy, removing the confounder of the heterogeneity of the disease and unsurvivable injury, and to maximise the chance of a TBI clinical trial’s success.

## Data Availability

All data produced in the present study are available upon reasonable request to the authors.

## Abbreviations

EVD: External ventricular drain
GCS: Glasgow coma scale
GOS-E: Glasgow Outcome Scale-Extended
Iba1: ionized calcium-binding adaptor molecule 1
ICP: Intracranial pressure
IHC: Immunohistochemistry
IHP: Independent Healthcare Professional
INR: International normalized ratio
MAP2: Microtubule-associated protein-2
NeuN: Neuronal nuclear protein
OCT: Optimal cutting temperature
SHIBA: Severe Head Injury Brain Analysis
TBI: Traumatic brain injury
vWF: von Willebrand factor
YHU: Yip, Hasan and Uff

## Acknowledgements

The authors would like to thank the Neurosurgery Department, the Emergency Department and the Critical Care Department at the Royal London Hospital. We would also like to thank Hanane Trari Belhadef for laboratory assistance, and Prof. John Priestley and Prof. Rupert Pearse for critical reviewing of the manuscript.

